# Clinical and spatial characteristics of Severe Acute Respiratory Syndrome by COVID-19 in Indigenous of Brazil

**DOI:** 10.1101/2020.10.24.20218701

**Authors:** Daniele Melo Sardinha, Karla Valéria Batista Lima, Ana Lúcia da Silva Ferreira, Juliana Conceição Dias Garcez, Thalyta Mariany Rêgo Lopes Ueno, Yan Corrêa Rodrigues, Anderson Lineu Siqueira dos Santos, Rosane do Socorro Pompeu de Loiola, Ricardo José de Paula Souza e Guimarães, Luana Nepomuceno Gondim Costa Lima

**Affiliations:** Post-graduate Program in Epidemiology and Health Surveillance, Institute Evandro Chagas (PPGEVS/IEC). Ananindeua, PA, Brazil; Post-graduate Program in Parasitical Biology in the Amazon, Pará State University and Institute Evandro Chagas (PPGBPA/UEPA/IEC). Belém, PA, Brazil; Secretariat of State for Public Health of Pará (SESPA). Belém, PA, Brazil

**Keywords:** Severe Acute Respiratory Syndrome, Indigenous People, SARS-CoV-2, COVID-19, Brazil

## Abstract

The new coronavirus (SARS-CoV-2) emerged in Wuhan in China in December 2019, causing severe pneumonias and deaths, soon in March 2020 it reached pandemic level, affecting several countries including Brazil. The disease was named COVID-19, with characteristics of most infected having mild and moderate symptoms and a part severe symptom. The disease has already reached 158 ethnic groups, which have high vulnerability and limited access to health services. The objective is to investigate the clinical and spatial characteristics of Severe Acute Respiratory Syndrome of COVID-19 in the indigenous peoples of Brazil. It is an epidemiological, cross-sectional, analytical ecological study, based on data from the OpenDataSUS platform from 01/01/2020 to 31/08/2020. Profile variables, signs and symptoms and risk factors/comorbidities. The data were analyzed by Bioestat 5.3. There were 1,207 cases and 470 deaths. Profile: male gender (59.48%) means age 53 years. Signs and symptoms: fever (74.23%), cough (77.71%), sore throat (35.62%), dyspnea (69.34%), respiratory discomfort (62.80%), O2 saturation <95% (56.42%); and associated with mortality: dyspnea (80.0%) and O2 saturation <95% (69.36%). Risk factors and comorbidities (45.89%) were associated with deaths (54.04%). Comorbidities: Chronic Cardiovascular Disease (18.97%) and Diabetes Mellitus (18.97%), and associated with deaths: Chronic Cardiovascular Disease (24.46%). Being admitted to the ICU has a risk of death in (OR-3.96-<0.0001-CI-2,913/5,383) followed by not being vaccinated against influenza (OR-1.85-<0.0001-CI-1,358/2,528). The public and health policies of Brazil should be directed to control the dissemination of COVID-19 in this population, that COVID-19 evolves in the same intensity, however, the indigenous have vulnerabilities that can increase the impact of the pan-demic in this population.

## 1. Introduction

Severe Acute Respiratory Syndrome (SARS) is a severe respiratory illness characterized by the presence of flu symptoms associated with dyspnea or respiratory discomfort or persistent pressure in the chest or oxygen saturation (O2) <95% or facial cyanosis [1]. It is caused by several etiologies such as (influenza A, dengue, respiratory syncytial virus, adenovirus, hantavirus, and coronavirus), and other agents (pneumococci, other bacteria, Legionella sp., leptospirosis, etc.) [2].

Regarding viral etiologies, they are capable of causing major epidemics in the world, such as those caused by two coronaviruses, SARS-CoV characterized in 2003 by SARS in China, which spread to 29 countries and regions, and MERS-CoV (Middle East Respiratory Syndrome) in 2012 in Saudi Arabia. The transmission of both was through respiratory droplets directly and indirectly and most of the cases proved to be mild [3].

Coronaviruses are single tape RNA viruses with envelope. The glycoprotein (S) peaks around the spherical verion give the virus its characteristic “halo appearance”, i.e., a crown in electron microscopy. Six subgroups of coronavirus are human pathogens, the subgroup α coronavirus includes 229E and NL229E and the subgroup β coronavirus includes OC43, HKU1, SARS-CoV, and MERS-CoV [4]. The pathophysiological mechanism in SARS-CoV and MERS-CoV is performed by binding in the angiotensin-converting en-zyme 2 (ACE2) and is expressed in many human organ tissues, being highly expressed in the lungs, heart, and small intestine, however, the presence of ACE2 may not be the only requirement for tropism [5,6].

In December 2019, cases of severe pneumonia of unknown etiology appeared in Wuhan, China, with the association of the cases to a market in the region. In January 2020 the infectious agent was isolated, being of the type coronavirus. Thus, it was named SARS-CoV-2 for its similarity to SARS-CoV and the disease named COVID-19, also presenting several other similarities such as the pathophysiological and transmissibility mechanism [7]. In March 2020 the World Health Organization (WHO) declared the out-break of SARS-CoV-2 as a pandemic, the first case confirmed in Brazil in February 2020[8,9].

The literature cites risk factors for severity at COVID-19 such as chronic disease carriers, the elderly, obese and pregnant women, generally severe cases require intensive care and longer hospital stay and high mortality [10,11]. However, studies do not show the impact and characteristics of COVID-19 on the indigenous population.

The epidemiological surveillance of COVID-19 in Brazil is performed from two online platforms: E-SUS notifies and SIVEP-GRIPE. E-SUS notifies is an exclusive platform for suspected and confirmed cases of influenza syndrome (IS) by COVID-19 and SIVEP-GRIPE is a platform that performs surveillance of SARS by all etiologies, among them COVID-19, so SIVEP-GRIPE notifies any case of hospitalized SARS or death by SARS regardless of hospitalization [1].

In Brazil, according to COVID-19’s monitoring platform on Indigenous Peoples, based on data from the Articulation of Indigenous Peoples of Brazil (APIB), it shows that on 20/09/2020 it already had 32,615 cases on Indigenous Peoples, 818 deaths, and 158 affected peoples [12]. Another agency that performs surveillance of cases in indigenous people in Brazil is the Special Secretariat of Indigenous Health (SESAI), which reported the occurrence of 26,723 cases and 426 deaths, with a mortality rate of 1.59% [13]. The difference in information is observed, in the deaths is double, this shows the underreporting of cases in this population, as well as the ignorance of this disease in this group.

It stands out for the indigenous peoples, their characteristics with the worst human development rates. They have precarious and limited access to health services, high infant mortality rates, and the main diseases that affect them are tuberculosis, verminous, diar-rhea, and respiratory infections. It is also worth mentioning that institutional racism and the continuous loss of their land also result in food quality and lack of assistance. Because of the preservation of biodiversity and culture, these people live in rural and distant places, which increases vulnerability, where public policies do not yet reach associated with extreme poverty [14].

Due to the vulnerability of indigenous peoples and the difference in information be-tween APIB and SESAI, and the importance of knowing the clinical characteristics and factors associated with mortality, which becomes crucial for the development of intervention and prevention strategies. In this perspective, this study aims to investigate the clinical and spatial characteristics of severe acute respiratory syndrome by COVID-19 in indigenous Brazil.

## 2. Materials and Methods

The cross-sectional, analytical, ecological and epidemiological study, quantitative, of secondary base data, from data made available by the OpenDataSUS platform (https://opendatasus.saude.gov.br/) of the Ministry of Health of Brazil [15], regarding the surveillance data of Severe Acute Respiratory Syndromes (SARS), from the platform of In-formation System for Epidemiological Surveillance of Influenza (SIVEP-GRIPE) corresponding to the period from 01/01/2020 until 31/08/2020 of the notifications. (https://sivepgripe.saude.gov.br/sivepgripe/login.html?1).

The bank was obtained from the OpenDataSUS platform in Excel 2019 format, on the date of 09/05/2020, with the last update on 08-31/2020, the bank refers to SARS cases from 01/01/2020 to 08-31/2020. The SIVEP-GRIPE notification form is composed of 80 variables, referring to socio-demographic and clinical-epidemiological data (https://opendatasus.saude.gov.br/dataset/ae90fa8f3e94467ea33f94adbb66edf8/resource/54a46c6d-e0b5-40b7-8b74-85450d22ace3/download/ficha-srag-final-27.07.2020_final.pdf). The variables extracted according to the form were: date of notification (item 1), sex (item 8), municipality of residence (item 9), age (item 10), signs and symptoms (item 35), has risk/comorbidities factors (item 36), took flu vaccine (item 37), hospitalized in an intensive care unit (item 47), final classification: 5-SARS by COVID-19 (item 72) and evolution:2-death (item 74).

The inclusion criteria were notifications of residents in Brazil, the indigenous race, and final SARS classification by COVID-19. Blank notifications were excluded. The data were organized in excel spreadsheet 2019 and the analysis was performed by the statistical program Statistical Package for the Social Sciences 25.0 (SPSS-https://www.ibm.com/analytics/spss-statistics-software). From the chi-square, statistical tests for independence test and G test (Contingency Table L x C) at the same values or <0.05, to associate the significant variables. The results were presented in tables.

The binary logistic regression test was applied to evaluate the dependence of the variable death to the covariates admitted to an Intensive Care Unit (ICU) and unvaccinated against influenza at risk of death. For all tests, the alpha significance level of 0.05 was considered.

The Brazilian official territorial division (states, regions and municipalities) were obtained on the website of the Brazilian Institute of Geography and Statistics (IBGE) (https://www.ibge.gov.br/).

The geocodes from OpenDataSUS and IBGE were used to join Excel data to shapefile (Brazil municipalities). This shapefile (polygons) was converted to points (centroid - “Centers Inside”) using the polygon to point function of ET GeoWizards (https://www.ian-ko.com/). The shapefile (points) was used to performed the time series for months based on the date of first symptoms.

From these shapefiles, were generated: choropleth maps for the analysis of spatial and temporal distribution to spatially visualize the location of the deaths by COVID-19; and spatial scanning map (Scan) to identify spatial and temporal clusters with statistical significance. The Scan used the Bernoulli model (purely spatial analysis scanning for clusters with high rates) based on the cases (deaths) and control (lifes) of cases of COVID-19 in the indigenous peoples of Brazil.

Data processing, interpretation, visualization, temporal and spatial analysis were performed in ArcGis (https://www.arcgis.com/), SaTScan (https://www.satscan.org/), and TerraView software (http://www.dpi.inpe.br/terralib5/wiki/doku.php).

The data of this study were made publicly available, not containing personal data of patients such as name, address, and telephone contact, thus not presenting risks to the participants of the research, as well as being dispensed with the ethical opinion. This study is by Law No. 12,527 of 18/11/2011 (Law of Access to Information) [16].

## 3. Results

The database (DB) shows a total of 670,929 notifications, including negative ones, awaiting results and confirmed ones: (1) SARS due to influenza, (2) SARS due to another respiratory virus, (3) SARS due to another etiologic agent, (4) SARS unspecified and (5) SARS due to COVID-19. Thus, the indigenous race was filtered in the DB which resulted in 1,991 notifications with those confirmed for COVID-19, resulting in 1,207 notifications. Table 1 shows the profile of the indigenous people about sex and age group, being most of the cases male (718; 59.48% p<0.0001), predominating also the male sex in the deaths (314; 66.80% p<0.0001). About the age group, the average of the cases was 53 years, over the survivors the majority was between 21 and 50 years; already about the deaths, it was distinguished the >60 years (309; 65.74% p<0.0001) with the average of 64 years. Between the ages of 0 and 20 years there was no statistical difference between survivors and deceased, however, in the age group 51 to 60 years there was a significant difference between survivors and deceased (121;16,41% p<0.0001)..

**Table 1.**
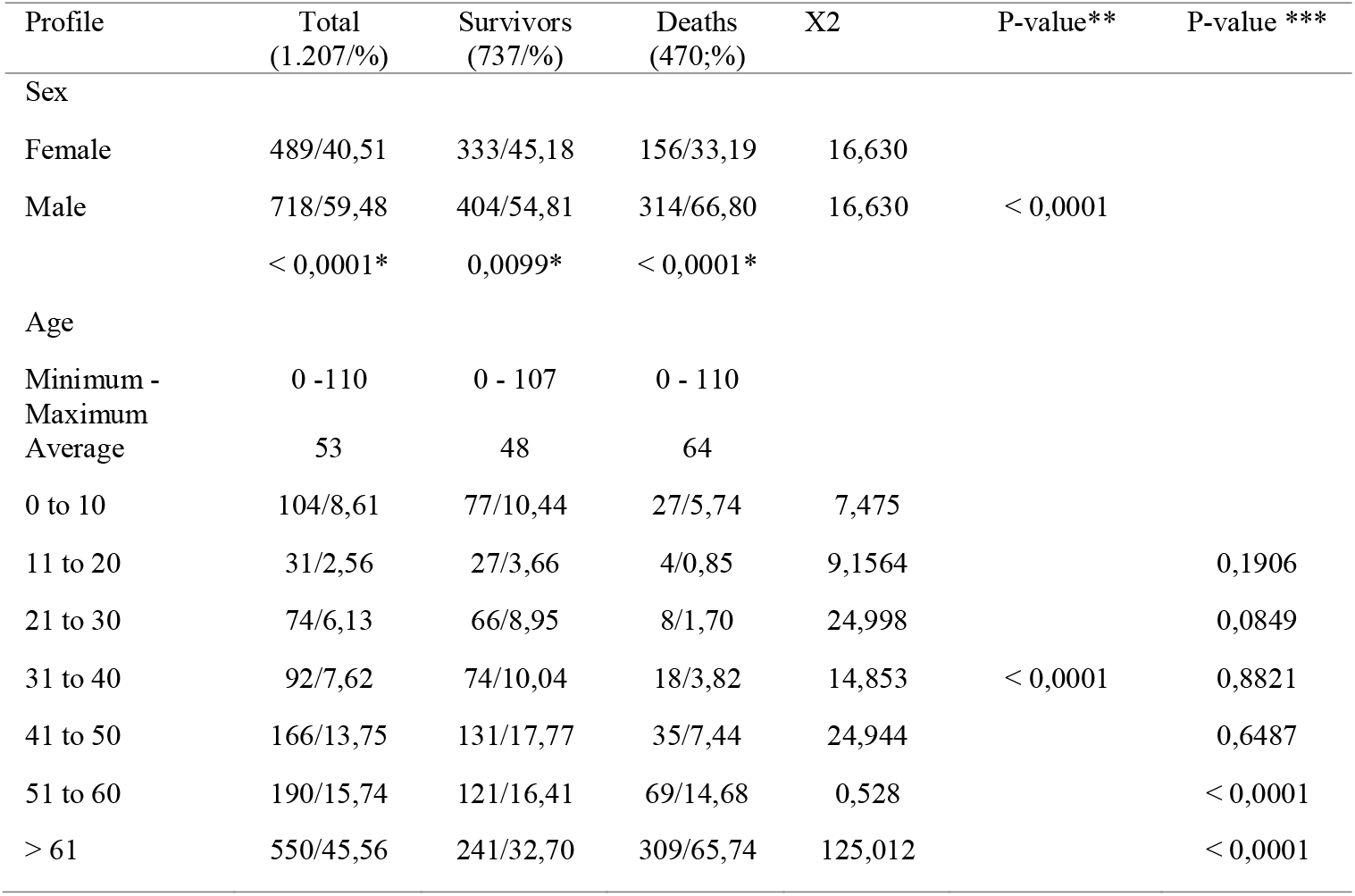
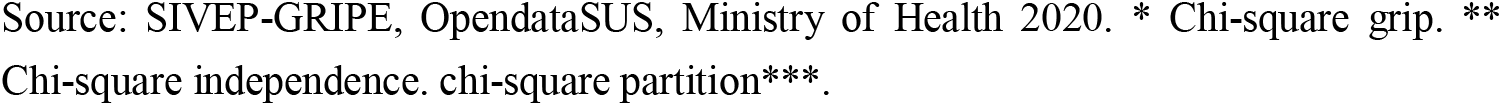
Profile of Severe Acute Respiratory Syndrome by COVID-19 in Indigenous Brazil, 2020.

| Profile | Total<br>(1.207/%) | Survivors<br>(737/%) | Deaths<br>(470/%) | X2 | P-value** | P-value *** |
| --- | --- | --- | --- | --- | --- | --- |
| Sex |  |  |  |  |  |  |
| Female | 489/40,51 | 333/45,18 | 156/33,19 | 16,630 |  |  |
| Male | 718/59,48 | 404/54,81 | 314/66,80 | 16,630 | < 0,0001 |  |
|  | < 0,0001* | 0,0099* | < 0,0001* |  |  |  |
| Age |  |  |  |  |  |  |
| Minimum -<br>Maximum | 0 - 110 | 0 - 107 | 0 - 110 |  |  |  |
| Average | 53 | 48 | 64 |  |  |  |
| 0 to 10 | 104/8,61 | 77/10,44 | 27/5,74 | 7,475 |  |  |
| 11 to 20 | 31/2,56 | 27/3,66 | 4/0,85 | 9,1564 |  | 0,1906 |
| 21 to 30 | 74/6,13 | 66/8,95 | 8/1,70 | 24,998 |  | 0,0849 |
| 31 to 40 | 92/7,62 | 74/10,04 | 18/3,82 | 14,853 | < 0,0001 | 0,8821 |
| 41 to 50 | 166/13,75 | 131/17,77 | 35/7,44 | 24,944 |  | 0,6487 |
| 51 to 60 | 190/15,74 | 121/16,41 | 69/14,68 | 0,528 |  | < 0,0001 |
| > 61 | 550/45,56 | 241/32,70 | 309/65,74 | 125,012 |  | < 0,0001 |

Source: SIVEP-GRIPE, OpendataSUS, Ministry of Health 2020. \* Chi-square grip. \*\* Chi-square independence. chi-square partition\*\*\*.

The clinical characteristics (Table 2) regarding the most evident signs and symptoms were: Fever (896:74.23%), Cough (938:77.71%,) Sore throat (430:35.62%), Dyspnea (837:69.34%), Respiratory Discomfort (758:62.80%), O2 Saturation <95% (681:56.42%). The signs and symptoms associated with mortality were dyspnea (376:80.0% p<0.0001) and O2 saturation <95% (326:69.36%) p<0.0001). The presence of risk factors and comorbidities (554:45.89%) was also associated with deaths (253:54.04% p<0.0001). The most evident comorbidities were: Chronic Cardiovascular Disease (229:18.97%) and Diabetes Mellitus (229:18.97%). The only comorbidity associated with deaths was Chronic Cardiovascular Disease (115:24.46% p<0.0002). Those vaccinated for influenza showed significance in survivors (193:26.18% p<0.0001).

**Table 2.** Clinical Characteristics of Severe Acute Respiratory Syndrome by COVID-19 in Indigenous Brazil, 2020.

| Clinical Characteristics | Total<br>(1.207/%) | Survivors<br>(737/%) | Deaths<br>(470/%) | X<sup>2</sup> | P-value<br>(survivors vs. deaths) |
| --- | --- | --- | --- | --- | --- |
| Fever | 896/74,23 | 551/74,76 | 345/73,40 | 0,210 | 0,6465 |
| Cough | 938/77,71 | 568/77,06 | 370/78,72 | 0,363 | 0,5469 |
| Sore Throat | 430/35,62 | 262/35,54 | 168/35,74 | 0,000 | 0,9941 |
| Dyspnea | 837/69,34 | 461/62,55 | 376/80,0 | 40,288 | < 0,0001 |
| Respiratory discomfort | 758/62,80 | 435/59,02 | 323/68,72 | 11,148 | 0,0008 |
| O<sub>2</sub> saturation <95% | 681/56,42 | 355/48,16 | 326/69,36 | 51,567 | < 0,0001 |
| Diarrhea | 177/14,66 | 108/14,65 | 69/14,68 | 0,005 | 0,9437 |
| Vomit | 117/9,69 | 74/10,04 | 43/9,14 | 0,169 | 0,6812 |
| Altered chest X-ray | 441/36,53 | 266/36,09 | 175/37,23 | 0,116 | 0,7336 |
| Other | 338/28,00 | 220/29,85 | 118/25,10 | 2,973 | 0,0847 |
| Risk Factors and Comorbidities | 554/45,89 | 301/40,84 | 253/54,04 | 18,978 | < 0,0001 |
| Pregnant | 35/2,89 | 35/4,74 | 0/0,00 | 30,3464 | < 0,0001 |
| Puerpera | 11/0,91 | 10/1,35 | 1/0,21 | 3,4667 | 0,0626 |
| Chronic Cardiovascular Disease | 229/18,97 | 114/15,46 | 115/24,46 | 14,541 | < 0,0002 |
| Chronic Hematological Disease | 8/0,66 | 4/0,54 | 4/0,85 | 0,0774 | 0,7809 |
| Down syndrome | 4/0,33 | 0/0,00 | 4/0,85 | 4,0945 | 0,0430 |
| Chronic Liver Disease | 9/0,74 | 2/0,27 | 7/1,48 | 4,1240 | 0,0423 |
| Asthma | 23/1,90 | 13/1,76 | 10/2,12 | 0,055 | 0,8143 |
| Diabetes Mellitus | 229/18,97 | 120/16,28 | 109/23,19 | 8,468 | 0,0036 |
| Chronic Neurological Disease | 14/1,15 | 5/0,67 | 9/1,91 | 2,7326 | 0,0983 |
| Another Chronic Pneumopathy | 24/1,98 | 10/1,35 | 14/2,97 | 3,086 | 0,0790 |
| Immunodepression | 18/1,49 | 7/0,94 | 11/2,34 | 2,891 | 0,0891 |
| Chronic Kidney Disease | 41/3,39 | 16/2,17 | 25/5,31 | 7,735 | 0,0054 |
| Obesity | 27/2,23 | 15/2,03 | 12/2,55 | 0,155 | 0,6938 |
| Other | 199/16,48 | 113/15,33 | 86/18,29 | 1,624 | 0,2026 |
| Influenza vaccination | 264/21,87 | 193/26,18 | 71/15,10 | 19,977 | < 0,0001 |

Source: SIVEP-GRIPE, OpendataSUS, Ministry of Health 2020.

In table 3, see the elements that were included in the binary logistic regression, so the dependent variable was death, and the number of deaths was associated with the outcome ICU hospitalized and not vaccinated against influenza.

**Table 3.** List of elements included in binary logistic regression.

| Case Processing Summary |  |  |  |
| --- | --- | --- | --- |
|  |  | N | Marginal Percentage % |
| Deaths (dependent variable) | Not | 737 | 61,1 |
|  | Yes | 470 | 38,9 |
| ICU | Not | 978 | 81,0 |
|  | Yes | 229 | 19,0 |
| Influenza vaccine | Yes | 264 | 21,9 |
|  | Not | 943 | 78,1 |
| Total |  | 1207 |  |

Source: SIVEP-GRIPE, OpendataSUS, Ministry of Health 2020.

Binary logistic regression showed that being admitted to the ICU and not being vaccinated against influenza are risk-to-death predictors. Thus, it was observed that being hospitalized in the ICU the risk of death due to odds 3.96 (OR 3,960: CI 2,913-5,383: <0.0001) followed by not being vaccinated against influenza due to odds 1.85 (OR 1,1853: CI 1,358-2,528: <0.0001). See table 4.

**Table 4.** Association of ICU interns and not vaccinated against influenza at risk of death in COVID-19 Severe Acute Respiratory Syndrome in indigenous peoples in Brazil, 2020.

| Variables | Test | df | p-value. | Odds ration | 95% *C.I. for Odds ratio |  |
| --- | --- | --- | --- | --- | --- | --- |
|  |  |  |  |  | Inferior | Upper |
| ICU patients | 77,196 | 1 | < 0.0001 | 3,960 | 2,913 | 5,383 |
| Not vaccinated against influenza | 15,157 | 1 | < 0.0001 | 1,853 | 1,358 | 2,528 |
| Constant | 29,534 | 1 | < 0.0001 |  |  |  |

Source: SIVEP-GRIPE, OpendataSUS, Ministry of Health 2020. \* Confidence Interval.

In the spatial analysis we showed the cases per month, and observed that the cases and clusters were concentrated in the northern region of Brazil, where the indigenous population and villages are larger because they are located in the Brazilian Amazon. (figure 2).

**Figure 1.**
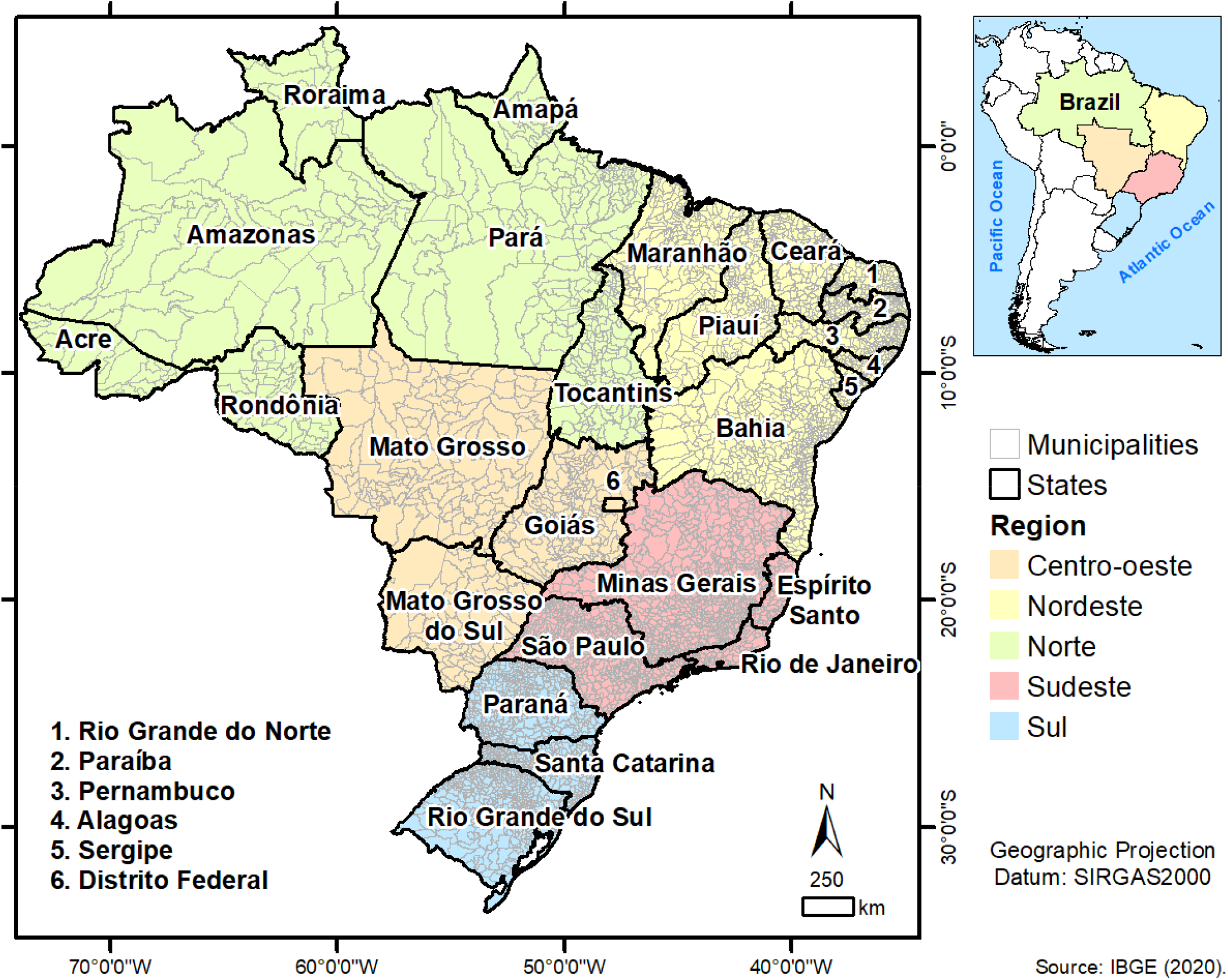
Shows the localization of Brazil and the Brazilian official territorial division.

**Figure 2.**
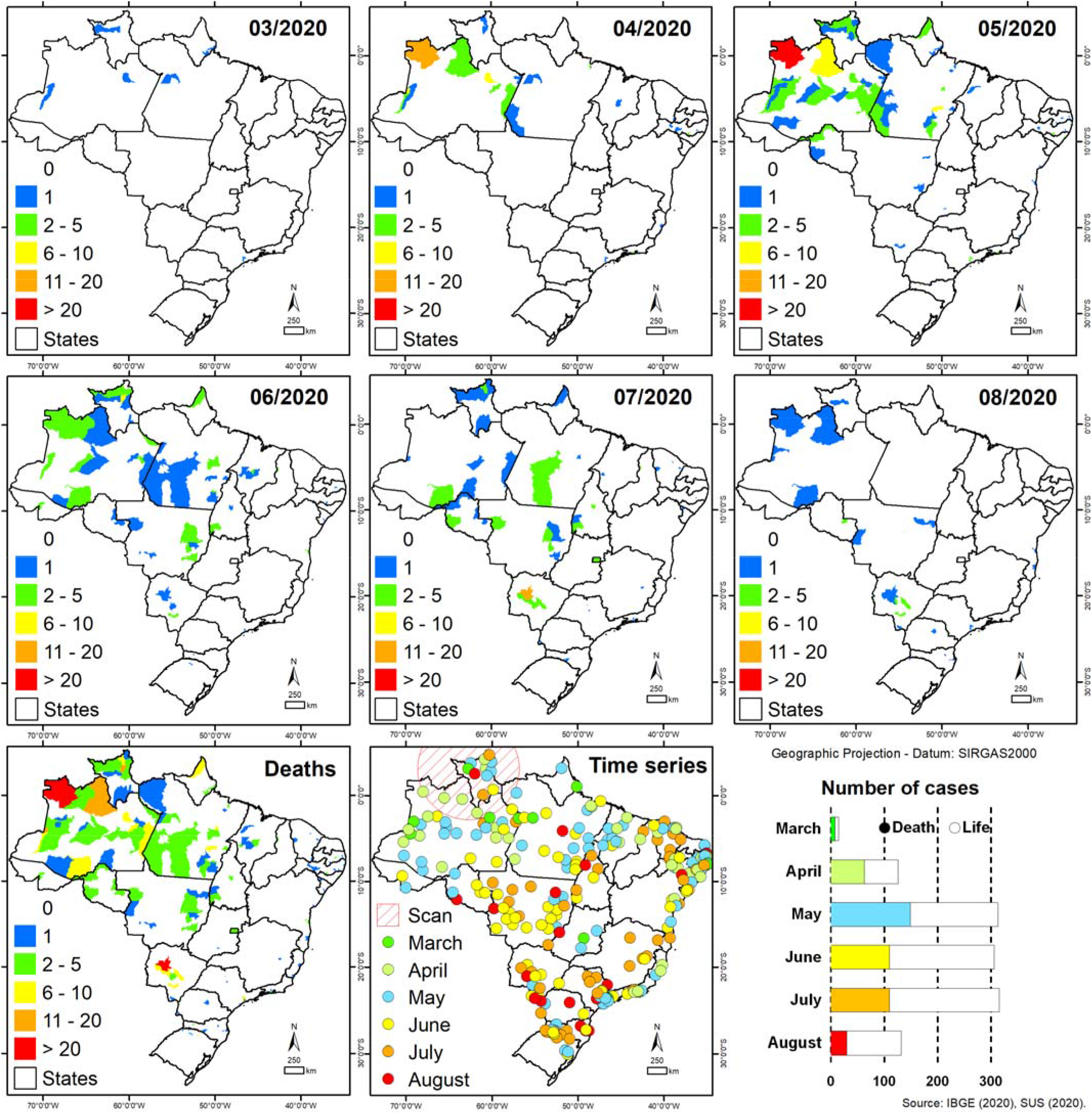
Shows the choroplethic maps of COVID-19 deaths per month (March to August) and for the entire study period (Deaths), and the time series of cases by COVID-19 per month.

## 4. Discussion

This study highlighted the clinical characteristics of SARS in indigenous Brazilians with an average age of 53 years (59.48%), being similar to a study [17] in non-indigenous patients hospitalized for SARS/COVID-19, however, the average age was 62 years, but approaching the results of two other studies with patients hospitalized for SARS by COVID-19 in non-indigenous patients, in one the average was 55 years, and in the other 50 years [18,19]. Therefore, there are no discrepancies between the average age and sex, between indigenous and non-indigenous people in SARS by COVID-19.

Concerning sex, it was significant in the deaths of the male (66.80%) with a mean age of 64 years. A large study with 17,278,392 cases, and concerning the deaths that were 10,926, identified the male gender and higher age as a predictor for mortality by COVID-19[20]. Another survey compared the deaths by COVID-19 from Italy 1,625 and China 1,023, and both were significant in individuals over 70 years, however, the lethality in Italy was higher (7.2%) because the infected were older, already in China was (2.3%) [21]. In these studies, cited were not specifically with indigenous people, but it is observed the similarity in the variables associated with death by COVID-19.

The most evident signs and symptoms in this study were fever (74.23%), cough (77.71%,) sore throat 35.62%), dyspnea (69.34%), respiratory discomfort (62.80%), O2 saturation <95% (56.42%). These results were similar in several studies [22–25], showing the classical picture of SARS associated with pulmonary involvement, evidenced by imaging examination, such as chest tomography [26]. This was also evident in this study the radiographic alterations in 36.53% of the cases. The Brazilian Ministry of Health established criteria for the definition of clinical diagnosis-imaging, in the impossibility of performing the molecular test or epidemiological link, and thus associated with the clinical characteristics the tomography should evidence by one of these changes: opaqueness in bilateral, peripheral frosted glass, with or without consolidation or visible intralobular lines (“paving”), or opaqueness in the multifocal frosted glass of rounded morphology with or with-out consolidation or visible intralobular lines (“paving”), or sign of reverse halo or other findings of pneumonia in an organization (observed later in the disease)[1]. Regarding deaths, the associated signs and symptoms were dyspnea (80.0%) and O2 saturation <95% (69.36%). Dyspnea and O2 saturation less than 95% indicate severe pulmonary involvement, which requires intensive care and mechanical ventilation if O2 saturation does not reach up to 92% with noninvasive oxygen therapy [27]. Studies relate the pulmonary involvement of other organs such as the heart to a storm of inflammatory cytokines, characterized by a sudden acute increase in circulating levels of different pro-inflammatory cytokines, IL-6, IL-1, TNF-α, and interferon, caused by the activation of several immune cells such as macrophages, neutrophils and T cells from the circulation to the site of infection causing damage to the vascular barrier, capillary damage, diffuse alveolar damage and multi-organ failure[28–30]. Thus, hyper inflammation influences O2 saturation drop and myocardial stability complications, causing the cardiopulmonary deficiency, with acute cardiac injury and lung injuries, which are serious complications directly associated with mortality [31,32].

The presence of risk factors and comorbidities (45.89%) of the cases was also evidenced, as well as associated with the deaths (54.04%). The most prevalent comorbidities were chronic heart diseases (18.97%) and diabetes (18.97%). However, the only significant comorbidities separately associated with deaths were chronic heart diseases (24.46%). Being corroborated by several types of research [33–35], However, the studies also showed not only heart disease as a factor of complications and deaths but also diabetes, obesity, and chronic lung disease, specifically the most significant comorbidity in these studies was hypertension and diabetes. A meta-analysis with 1,527 cases, showed that the prevalence of hypertension, diabetes, and cerebrovascular diseases, were evident two to three times more in patients hospitalized by COVID-19 in the Intensive Care Unit, than those who were in the ward, as they also highlighted the systemic complications, the most evident being the acute cardiac injury in 8%, directly associated to mortality [36]. In this way, the results meet the literature.

For the variable “to be vaccinated against the flu” there was significance in the survivors (26.18%). A research in Italy over 65 years old with high vaccination coverage of influenza, detected the association with a reduced spread and a less severe clinical expression of COVID-19[37]. A study highlights that mass immunization against influenza in the COVID-19 pandemic aims to minimize cases of co-infection that can be a factor of se-verity and mortality, as well as protecting risk groups that are similar in both diseases [38]. Thus, a similar result is shown in those vaccinated against influenza as a protection factor for severity of COVID-19, however, no studies are explaining this mechanism.

In the special distribution of cases per month, we observed the peak of cases in May 2020, similar to the peak of cases to the other groups, and the cluster of cases and deaths were significant in the northern region of Brazil, brazilian amazon region where it has the largest metropolitan indigenous population in the country, showing that COVID-19 reached the most distant villages of the metropolitan regions, similar to the results of Silva et al [39] who conducted a similar study however considering all etiologies of SRAG in indigenous peoples.

In Brazil, vaccination against COVID-19 began in January 2021, and indigenous peoples were priorities together with health professionals in the first stage of the vaccination campaign, as they are a highly vulnerable group, as well as a report showed that cases and deaths from COVID-19 decreased after the second dose of the vaccine in the state of Minas Gerais in Brazil [40].The limitation of this research is that there are no studies of SARS by COVID-19 in the indigenous population, and it is not possible to compare the variables, however, it was possible to compare the clinical characteristics with other ethnic groups or even general. As well, it was shown the similarity in the different population groups, showing that COVID-19 presents in the same intensity and needs attention in these groups as indigenous since these people have limited access to health services, as well as the prevalence of other diseases that can potentialize the complications of COVID-19.

## 5. Conclusions

It was possible to evidence the clinical characteristics of Severe Acute Respiratory Syndrome in Indigenous Brazil, from the data of SIVEP-GRIPE. They are mostly male, both in cases and deaths, in cases of adults and older deaths, the most common signs and symptoms being fever, cough, sore throat, dyspnea, respiratory discomfort, and O2 satu-ration <95%, however, the signs and symptoms associated with the deaths were dyspnea and O2 saturation <95%. The most prevalent comorbidities were chronic heart disease which was associated with deaths and diabetes.

It has also been shown that admission to the ICU and not being vaccinated against influenza, are factors that increase the chances of death.

It is emphasized that Brazil’s public and health policies are directed towards control-ling the spread of COVID-19 in the indigenous populations since it has been shown that the disease evolves in the same intensity in this group, however, the indigenous have vulnerabilities that can potentialize the impact of this pandemic on this population.

## Data Availability

data are available on the opendatasus platform of the Brazilian Ministry of health

## Footnotes

** Special description of the title. (dispensable)

